# Integrating multiparametric imaging and electrophysiology reveals thalamic pathology

**DOI:** 10.64898/2026.07.28.26359146

**Authors:** Roy A.M. Haast, Julia Makhalova, Lucas Gauer, Jacob Knight, Mohamed Mounir El-Mendili, Hugo Dary, Samuel Medina Villalon, Jean-Philippe Ranjeva, Wafaa Zaaraoui, Fabrice Bartolomei, Maxime Guye

**Affiliations:** Aix Marseille Univ, CNRS, CRMBM, Marseille, France; APHM, Hôpital Universitaire Timone, CEMEREM, Marseille, France; Department of Epileptology and Clinical Neurophysiology, APHM La Timone Hospital, Marseille, France; INS, INSERM UMR 1106, Aix-Marseille University, Marseille, France

**Keywords:** focal epilepsy, thalamus, ultra-high-field MRI, multiparametric MRI, intracranial electrophysiology

## Abstract

The thalamus is increasingly recognized as a key node within epileptogenic networks, yet how recurrent seizure activity shapes its tissue integrity remains poorly understood. Existing evidence has largely relied on individual structural, functional, or electrophysiological modalities, limiting an integrated understanding of thalamic pathology.

Here, we combined quantitative sodium, structural, and functional 7 Tesla MRI with stereotactic electroencephalography (SEEG) and clinical characterization to establish a multimodal framework for investigating thalamic involvement in drug-resistant focal epilepsy.

Multiparametric MRI revealed widespread increases in total sodium concentration, providing the first evidence of altered thalamic sodium homeostasis in focal epilepsy, together with focal elevations of the short T_2_* sodium signal fraction within lateral thalamic regions and increased regional homogeneity. Integrating these complementary measures identified a robust MRI profile that distinguished patients from controls and independently identified patients with SEEG-defined epileptogenic thalami. Reduced thalamic volume was associated with greater ictal thalamic recruitment, whereas multivariate behavioral analyses demonstrated that complementary MRI features differentially reflected epileptogenic network burden, disease chronicity, and demographic characteristics.

By integrating measurements spanning tissue pathology, intracranial electrophysiology, and clinical phenotype, this study establishes a framework for characterizing pathological network nodes in focal epilepsy. Such multimodal imaging profiles may support patient stratification and individualized therapeutic strategies, including epilepsy surgery and targeted neuromodulation.

## Introduction

Focal epilepsy is a chronic neurological condition marked by recurrent, spontaneous seizures arising from abnormal neuronal rhythms. Seizures spread from affected areas within the epileptogenic zone network (EZN) to connected areas within the propagation zone network (PZN)^1^. Although the path of epileptic seizure spreading across the brain is highly patient-specific, the thalamus is among the most observed areas in either the EZN or PZN^2^. Its central role as a hub of nuclei that integrate, modulate and relay information across the brain^3,4^ likely contributes both to its frequent recruitment during seizures and to its influence on epileptogenic network organization.

Given its central role in epileptogenic networks, there is growing interest in developing imaging biomarkers that characterize thalamic involvement in focal epilepsy. Such biomarkers could improve our understanding of disease mechanisms, facilitate patient stratification, and support individualized therapeutic strategies, including epilepsy surgery and deep brain stimulation (DBS) for drug-resistant focal epilepsy (DRFE) patients^5,6^. Accordingly, recent research has focused on identifying changes within the thalamus. Stereotactic electroencephalography (SEEG) recordings have revealed transient thalamic recruitment during seizures, marked by abnormal synchronization and excitability^7–11^. Notably, greater thalamic epileptogenicity is linked to more extensive epileptogenic networks and a higher likelihood of surgical treatment failure^2^. Structural and functional magnetic resonance imaging (MRI) studies have documented chronic changes, including thalamic atrophy^12–16^, altered longitudinal relaxation time T_1_ ^13^, disrupted thalamocortical connectivity^14,17^, and abnormal local activity measured via blood oxygen-level dependent (BOLD) functional MRI^18^. Although these studies consistently demonstrate thalamic abnormalities, they have largely examined individual imaging modalities in isolation. Consequently, it remains unclear whether these alterations reflect independent processes or complementary manifestations of a common thalamic pathological phenotype.

Characterizing the thalamic pathological phenotype requires imaging markers that probe complementary aspects of tissue physiology. Whereas conventional proton MRI provides sensitive measures of tissue structure and function, sodium (^23^Na) MRI offers a unique, non-invasive window into ionic homeostasis. Neuronal viability relies on tightly regulated intra- and extracellular Na^+^ concentrations that support normal neuronal signaling, and disruption of this balance can impair cellular function and viability^19–21^. Accordingly, sodium MRI has emerged as a non-invasive method for assessing brain health *in vivo*^22^. In patients with DRFE, previous studies have reported globally increased total sodium concentration (TSC) together with more localized elevations of the short T_2_* sodium fraction (*f_Na_*) within cortical epileptogenic regions^21,23^. Because *f_Na_* is thought to reflect tissue excitability and, potentially, epileptogenicity^23^, sodium MRI provides information that complements conventional structural and functional imaging. Whether sodium dysregulation also characterizes the thalamus, and how it relates to established structural and functional abnormalities, remains unknown. More broadly, it remains unclear whether these diverse imaging abnormalities converge into a coherent thalamic phenotype that reflects electrophysiological involvement in epileptogenic networks.

Here, we developed a multimodal framework that combines multiparametric quantitative ^23^Na and ^1^H 7 Tesla (7T) MRI with intracranial SEEG to characterize thalamic involvement in well-characterized patients with DRFE. Quantitative MRI measures of sodium homeostasis (TSC and *f_Na_*), intrinsic functional organization (regional homogeneity and fractional amplitude of low-frequency BOLD fluctuations), and structural integrity (volume and T_1_) were integrated into a multiparametric MRI profile and related to electrophysiological measures of thalamic epileptogenicity and clinical network characteristics using multivariate, mediation, and machine-learning analyses. This framework may facilitate the development of non-invasive biomarkers to improve patient stratification and support individualized therapeutic decision-making, including epilepsy surgery and DBS.

## Materials and methods

### Dataset

#### Participants

We studied consecutive patients diagnosed with DRFE who underwent pre-surgical evaluations at our facility from June 2017 to October 2020, including 7T brain MRI and SEEG recordings. The 7T MRI scans were collected within the scope of two prospective studies focused on pre-surgical evaluations for DRFE: RHU EPINOV clinical trial (NCT03643016) and NEURO7T (2016-A01785-46). The SEEG recordings were systematically performed for all patients as part of routine clinical care following the French guidelines^24^. We also recruited age- and sex-matched healthy controls for comparison of the imaging data. Both studies received approval from the local ethics committee (*Comité de Protection des Personnes sud Méditerranée 1*), and all participants provided written informed consent, in compliance with the Declaration of Helsinki. This study complies with the requirements of the national personal data regulation committee (*Commission Nationale de l’Informatique et des Libertés*).

#### 7T MRI acquisition

All MRI data were acquired on a Siemens Magnetom 7T Step 2 MRI system (Siemens Healthineers, Erlangen, Germany). For each subject, ^1^H MRI B_1_ ^+^, T_1_ and resting-state functional MRI (rs-fMRI), and multi-echo ^23^Na MRI data were collected using a ^1^H 1Tx/32Rx (Nova Medical, Wilmington, USA) and dual-tuned ^23^Na/^1^H QED birdcage coils, respectively^23^.

#### Structural imaging

Whole brain B_1_^+^ and T_1_ maps were acquired using a spin echo-based sequence, assessing the ratio of consecutive spin and stimulated echoes^25^, and the 3D Magnetization Prepared with 2 Rapid Acquisition Gradient Echoes (MP2RAGE) sequence, respectively^26^. Acquisition parameters for the MP2RAGE sequence were as follows: TR_mpr_/TR/TE/TI_1_/TI_2_ = 5000/7.4/3.13/2750 ms, α_1_/ α_2_ = 6/5°, FOV = 240 mm (matrix size: 402 ⨉ 402), 256 partitions, a parallel imaging acceleration factor of 3 (2D GRAPPA), and Partial Fourier factors of 6/8 in both phase- and partition-encoding directions. This resulted in an isotropic nominal voxel size = 0.6 mm and a total acquisition time of 10.12 minutes.

#### Sodium imaging

Whole-brain sodium MRI data with a 3 mm isotropic nominal voxel size were acquired with a multi-echo density-adapted 3D projection reconstruction pulse sequence^27,28^. Acquisition parameters included: TR = 120 ms, 5000 spokes and 384 radial samples per spoke. The sequence was repeated three times in the same session to obtain 24 TEs ranging from 0.2 ms to 70.78 ms: 1^st^ acquisition: 0.20, 9.70, 19.20, 28.70, 38.20, 47.70, 57.20, 66.70 ms, 2^nd^ acquisition: 1.56, 11.06, 20.56, 30.06, 39.56, 49.06, 58.56, 68.06 ms, 3^rd^ acquisition: 4.28, 13.78, 23.28, 32.78, 42.28, 51.78, 61.28, 70.78 ms, with a total acquisition time of 30 minutes. Six tubes with known sodium concentrations (from 25 to 100 mmol/L within 2% of agar gel) were placed within the field of view as reference for quantification^28^.

#### Functional imaging

Human Connectome Project-style whole-brain resting-state functional MRI (rs-fMRI) data with an 1.6 mm isotropic nominal voxel size were acquired using a 2D Multi-Band (MB) Echo Planar Imaging (EPI) sequence to perform timeseries analysis^29^. Acquisition parameters were: TR/TE = 1000/22 ms, α = 38°, FOV = 208 mm (matrix size: 130 ⨉ 130), 85 partitions, a parallel imaging acceleration factor of 2 (2D GRAPPA), MB factor of 5, Partial Fourier factors of 7/8 in the phase-encoding direction, and total acquisition time of 10.31 minutes. An additional rs-fMRI dataset with reverse-phase encoding (left-right instead of right-left) but otherwise identical scanning parameters was acquired to correct the functional data for EPI readout-related geometrical distortions.

#### Electrophysiological recordings

After the 7T MRI scans, and as part of the clinical care, multi-contact intracerebral electrodes (10-18 contacts, 2 mm length, 0.8 mm diameter, 1.5 mm apart, Alcis, France) were implanted in patients’ brains based on clinical hypotheses using a ROSA™ stereotactic surgical robot. A cranial CT scan confirmed the absence of complications and accurate electrode placement. Signals were recorded with a 256-channel Natus system, sampled at 512 Hz, and stored on a hard disk (16 bits/sample) without digital filtering. The acquisition included high-pass (0.16 Hz at -3 dB) and anti-aliasing low-pass (170 Hz at 512 Hz) filters.

### Data analysis

#### Thalamic segmentation, T1 and volume

B_1_^+^ corrected T_1_ data were processed as described previously, including the automatic segmentation of the thalamus and its nuclei using the 7TAMIBrain atlas^13,30^. Additional, probability-based brain tissue segmentation of cerebrospinal fluid (CSF), white-matter (WM) and gray-matter (GM) was performed on the brain-extracted T_1_w-UNIDEN images using FSL’s (v.5.0.6) *fast*^31^. Considering the lower resolution of the functional and sodium MRI data, thalamic nuclei were grouped into medial, posterior, lateral, and anterior segments to improve the reliability of our results (see Table 1). However, the results of the anterior segment remain to be interpreted with caution as it was captured by a limited number of voxels for participants.

**Table 1.** Thalamic segments.

| Segment | 7TAMIBrain atlas region-of-interest |
| --- | --- |
| *Lateral* | Lateral |
| *Medial* | Medial<br>Centromedian<br>Mediodorsal<br>Central-lateral<br>Habenula |
| *Posterior* | Lateral geniculate<br>Medial geniculate<br>Lateral posterior<br>Pulvinar anterior<br>Pulvinar |
| *Anterior* | Anterior |

#### Sodium homeostasis

We used the fitting procedure as described by Ridley *et al.* (2018)^28^ which has been widely applied in ^23^Na MRI to separate short and long components^27^.

#### Biexponential modeling of the ^23^Na T_2_* signal decay

For each voxel, the mean signal intensity across all 24 echo times (TEs) was extracted and fitted with a biexponential decay model of the form:

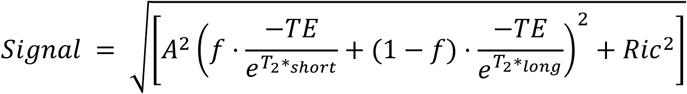

where *A* is a scaling factor, *f* represents the proportion of the short T_2_* component, and *Ric* refers to a Rician noise-related scaling parameter as described in Gudbjartsson and Patz (1995)^32^. This model yields voxel-wise estimates of the short and long T_2_* relaxation components (T_2_*_short_, T_2_*_long_) and their relative signal contributions. From the extrapolated intercepts of the decay curves, we computed the corresponding magnetization values: M_0,SF_ = *A*·*f* and M_0,LF_ = *A*·(1-*f*).

#### TSC and *f*_Na_ quantification

Since the fitted signal amplitudes reflect relative rather than absolute units, calibration was performed using the reference tubes included in the acquisition. For each tube, monoexponential fits of the multi-TE signal were used to estimate M_0_ and T_2_* values (MATLAB R2012a), yielding highly consistent fits across subjects (mean r^2^=0.97±0.01). A linear calibration curve was then generated between measured M_0_ values and the known sodium concentrations of the tubes, enabling conversion of the *in vivo* magnetization parameters M_0,SF_ and M_0,LF_ into quantitative sodium concentration estimates (Na_SF_, Na_LF_). TSC was then calculated by taking the sum of Na_SF_ and Na_LF_, while *f_Na_* was defined by the ratio between Na_SF_ and TSC (Fig. 1A).

**Fig. 1.**
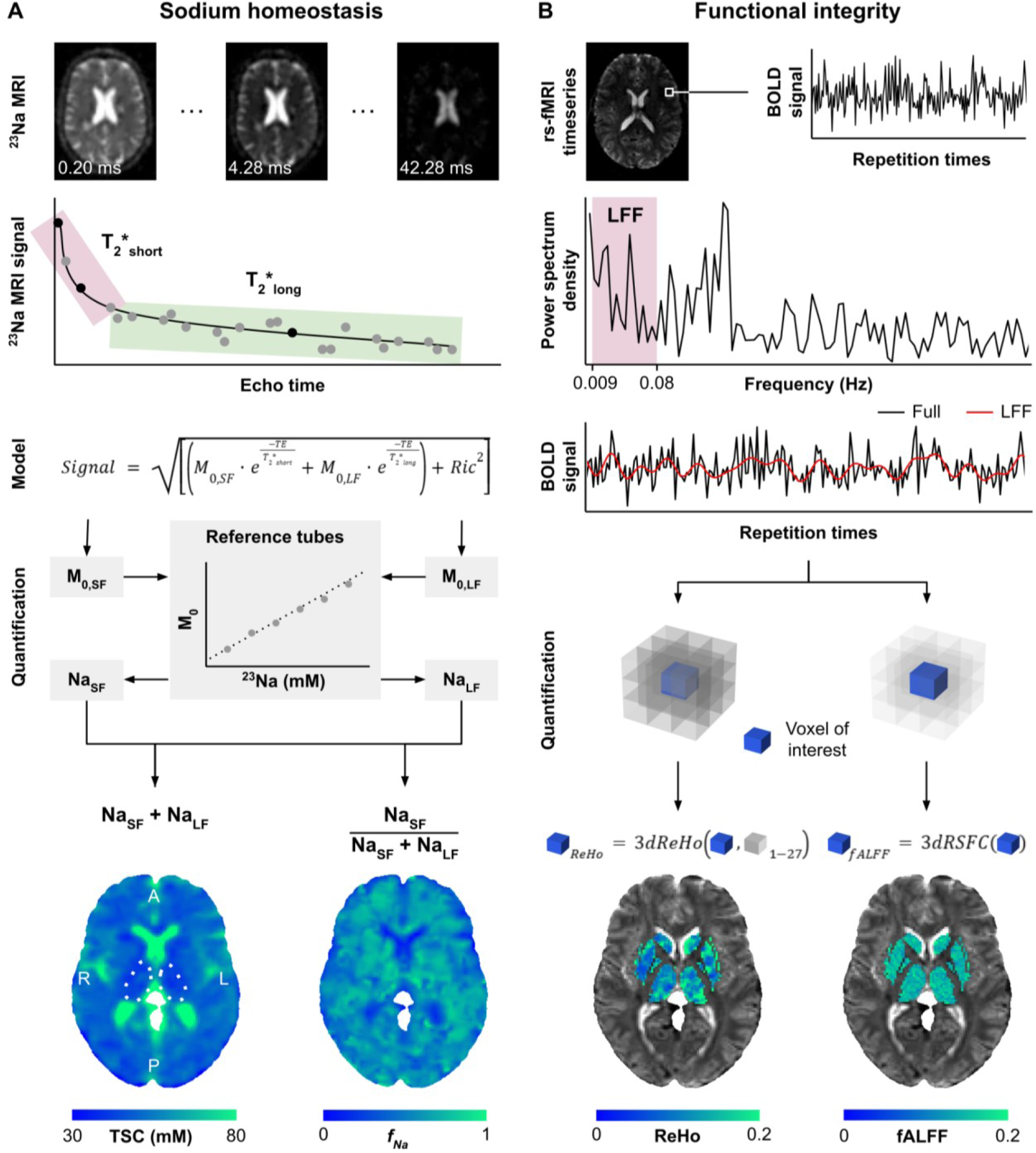
MRI-based assessments of thalamic sodium homeostasis and functional integrity. (A) Total sodium concentration (TSC, in mM) and short T_2_* fraction of the sodium signal (f_Na_; calculated as Na_SF_/TSC) maps were derived from ²³Na MRI using a biexponential signal decay model, with calibration based on reference phantoms containing known sodium concentrations. (B) Regional homogeneity (ReHo) and fractional amplitude of low-frequency fluctuations (fALFF) maps were computed from resting-state fMRI data using AFNI’s *3dReHo* and *3dRSFC* functions, respectively.

#### fALFF and ReHo

Functional integrity was assessed by calculating the fractional amplitude of low frequency fluctuations (fALFF) and regional homogeneity (ReHo) from preprocessed and denoised BOLD data^33,34^. ReHo characterizes the spatial coherence of intrinsic BOLD activity across neighboring voxels, whereas fALFF quantifies the relative strength of spontaneous low-frequency fluctuations within a given region.

#### rs-fMRI preprocessing

The preprocessed BOLD images were obtained using fMRIprep (v20.2.3)^35^, which is based on Nipype (v1.6.1)^36^. For each of the two rs-fMRI runs, a reference volume and its skull-stripped version were generated using a custom methodology of fMRIPrep. A B_0_ fieldmap was then estimated based on the two EPI references with opposing phase-encoding directions with AFNI’s (v16.2.07) *3dQwarp*^37^. Based on the estimated susceptibility distortion, a corrected EPI reference was calculated for co-registration with the anatomical T_1_w reference using FreeSurfer’s *bbregister* with six degrees of freedom^38^.

Head-motion parameters with respect to the BOLD reference (transformation matrices, and six corresponding rotation and translation parameters) were estimated before any spatiotemporal filtering using FSL’s *mcflirt*^39^. BOLD runs were slice-time corrected using AFNI’s *3dTshift*^37^. The BOLD time-series (including slice-timing correction) were resampled onto their original, native space by applying a single, composite transform using ANTs’ (v2.3.3) *antsApplyTransforms*, configured with Lanczos interpolation^40,41^, to correct for head-motion and susceptibility distortions. These resampled BOLD time-series will be referred to as preprocessed BOLD data.

Several confounding time-series were calculated based on the preprocessed BOLD: framewise displacement (FD) and three region-wise global signals. FD was calculated following the definitions by Power *et al.* (2014)^42^. The three global signals are extracted within the anatomically-derived eroded CSF and WM masks, and the whole-brain mask. The head-motion estimates calculated in the correction step were also placed within the corresponding confounds file. The confounds timeseries derived from head motion estimates were expanded with the inclusion of temporal derivatives and quadratic terms for each leading to a total of 24 motion parameters^43^.

#### Noise removal

Nilearn’s (v0.10.4) *clean_img* was then used to regress the confound timeseries from the preprocessed BOLD data simultaneously, after removal of the first five volumes. The “simple+gsr” strategy as defined by Wang *et al.* (2024)^44^ was followed by including the 24 motion parameters and the three global signals listed above. Note that scrubbing/censoring of high motion volumes was not performed for this study as this renders the temporal sampling irregular and prevents the application of a Fourier Transform to extract the required frequency spectrum for quantification of fALFF. Instead, mean framewise displacement (FD) across the whole timeseries was used to correct for motion differences across participants during the statistical analyses.

#### fALFF and ReHo calculation

The fALFF and ReHo parameter maps were computed using AFNI’s *3dRSFC* and *3dReHo* functions^45^. Briefly, *3dRSFC* first computed the amplitude of low frequency fluctuations (ALFF), represented by the sum of low frequency fluctuations spectral amplitudes of the Fourier-transformed denoised and filtered BOLD data^46^. The fALFF is then calculated by scaling ALFF by the sum of the time series’ amplitudes across the full spectrum (i.e., without prior band-pass filtering)^34(p20)^. Note that *3dRSFC* provided the band-pass (0.008-0.09 Hz) filtered timeseries that was used as input to *3dReHo* to calculate Kendall’s W for each voxel based on its concordance with its face-, edge-, and node-wise (i.e., 27) neighborhood voxels (Fig. 1B).

#### Epileptogenic zone

Two experienced neurologists (authors F.B. and J.M.) visually inspected signals from at least two SEEG-recorded electroclinical seizures per patient using AnyWave software (https://gitlab-dynamap.timone.univ-amu.fr/anywave)^47^, focusing on signals from contacts in the gray matter, excluding those with artifacts. The epileptogenicity index (EI) and connectivity EI (cEI) were calculated for each bipolar derivation. EI combines the ratio of fast frequencies (beta/gamma) relative to slower frequencies and the time of involvement of each region^48^. This was then combined with a directed connectivity measure to calculate cEI, to cope with seizure-onset patterns without low-voltage fast activity^49^. Derivations with an EI ≥ 0.4 and/or cEI ≥ 0.65 and corrected by visual inspection were classified as the epileptogenic zone network (EZN), as previously established^50^. In addition to EI and cEI, interictal epileptogenicity markers were quantified as well, including spike and ripple rates^50^.

In each patient, an automated anatomical localization and labeling of each electrode contact was conducted using the Gardel software (https://meg.univ-amu.fr/wiki/GARDEL) in the EpiTools software suite^51^. The pre-implantation T_1_-weighted MRI was co-registered with the post-implantation CT images, followed by automatic identification and anatomical localization of each electrode contact. Co-registration was then performed with the Virtual Epileptic Patient (VEP) atlas for automated brain parcellation (https://ins-amu.fr/vep-atlas)^52^, in conjunction with the 7TAMIbrainDGN atlas for automated parcellation of thalamic nuclei and basal ganglia^30^. Coordinates of bipolar derivations located within the thalamus, as well as region-of-interests defined by a 5 mm radius sphere around these coordinates^23^, were then transformed to the patient’s 7T MRI and MNI spaces by applying the transformation matrices derived from the linear coregistration between CT, MP2RAGE and MNI template images.

### Statistical analyses

Average TSC, *f*_Na_, fALFF, ReHo, T_1_, and volume were extracted for the left and right thalami and their lateral, medial, posterior and anterior segments. Note that TSC and *f_Na_* were weighted for gray matter-density to perform voxel-wise correction for the partial spread of neighboring structures (i.e., spilling effects). These multiparametric estimates were corrected for age, sex, hemisphere and estimated total intracranial volume effects based on the control data, using the *confounds* (v0.1.2) Python package^53^. Additional correction for mean FD was performed for the fALFF and ReHo metrics (see Supplementary Fig. 1).

#### Feature-wise comparison

Following z-scoring with respect to controls, assessment of statistical differences of MRI features between subject groups, segments, and side relative to seizure focus (i.e., ipsi-vs. contralateral) were explored using a mixed-effects model for repeated measures (with subject as random effect), followed by pairwise comparisons when necessary, Spearman’s rank correlation, and mediation analyses implemented in the *pingouin* (v0.5.3) Python package. In case of multiple comparisons, and if not stated explicitly otherwise, correction using Benjamini-Hochberg’s false discovery rate procedure was performed.

#### Partial least squares analysis

Partial least squares (PLS) analyses were carried out to further explore the variability in MRI-based features between groups (i.e., ‘mean-centered’ PLS) and with respect to the clinical features across patients (i.e., ‘behavioral’ PLS) using the *PyPLS* Python package^54,55^. Additional details concerning the PLS analyses can be found in Haast *et al.* (2023)^13^.

Imaging features were then selected by their absolute multivariate bootstrap ratio (BSR) values from the mean-centered PLS analysis for further analyses (i.e., automatic classification and composite MRI score). A BSR ≥ |2.58| was considered reliable, representing a 99% confidence interval^56^.

#### Automatic classification

Three classifier algorithms (logistic regression, linear support vector machine and random forest) were trained using *scikit-learn* (v1.6.1). Addressing sample size, Leave-One-Out Cross-Validation (LOOCV) was performed for each classifier, where models are trained on n−1 samples and evaluated on the held-out sample. Inside each training fold, class imbalance was addressed using the synthetic minority over-sampling technique^57^. Model hyperparameters were tuned using grid search with stratified 5-fold cross-validation on the training fold (n−2 samples). LOOCV predictions were then aggregated to compute overall ROC curves for each model. The same pipeline was implemented on a feature set of equal dimensions containing the least significant latent variables (i.e. those closest to zero) as per feature BSR. DeLong’s test was used to compare the ROC curves for each classifier when trained on the two distinct feature sets.

#### Composite MRI score

A patient-specific composite MRI score was calculated for each ipsilateral thalamus by projecting the selected MRI features onto the significant patient-versus-control PLS latent variable. Features were selected based on their bootstrapped ratios, while their corresponding PLS loadings were used as weights:

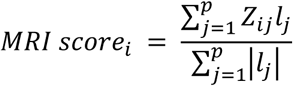

where *Z_ij_* denotes the control-referenced z-score of MRI feature *j* for patient *i*, *l_j_* is the corresponding PLS loading, and *p* is the number of selected ipsilateral MRI features. Scores were oriented such that higher values reflected stronger expression of the patient-like thalamic MRI signature. These scores were compared between patients with and without SEEG-defined thalamic epileptogenicity using a Mann–Whitney U test.

## Results

### Study cohort

A total of 30 patients with various EZN locations and 20 healthy controls were included in the final dataset (Table 2). Based on the anatomo-functional organization of the EZN, as derived from SEEG analysis, patients were categorized into distinct groups: (i) temporal lobe (TLE), where the EZN was confined to the temporal regions or had its primary epileptogenic focus in the temporal lobe with some extension to extratemporal areas (e.g., temporo-insular, temporo-frontal, temporo-occipital, N = 21) and (ii) non-temporal lobe (NTLE, N = 9). For the NTLE, the EZN affected the prefrontal regions (N = 3), parietal and/or occipital regions (N = 3), insula and/or opercular cortex (N = 2) or the primary motor and/or premotor cortex (N = 1). Owing to the limited number of patients with complete high-quality 7T MRI data, primary analyses compared all DRFE patients with healthy controls. Additional subgroup analyses (TLE, NTLE, controls) are presented in the Supplementary Data.

**Table 2.** Patient clinical characteristics.

| Demographics |  | Overall (N=30) | Patients with thalamic SEEG (N=22) |
| --- | --- | --- | --- |
| Age, mean [range] | - | 31.6 [15-57] | 32.8 [15-57] |
| Sex, n (%) | *Male* | 16 (53.3) | 8 (36.4) |
|  | *Female* | 14 (46.7) | 14 (63.6) |
| Clinical characteristics |  |  |  |
| Age at epilepsy onset, mean [range] | - | 15.4 [2-40] | 17.3 [2-40] |
| Disease duration, mean [range] | - | 16.9 [3-42] | 16.0 [3-42] |
| Epileptogenic zone lateralization, n (%) | *Left* | 14 (46.7) | 9 (40.9) |
|  | *Right* | 16 (53.3) | 13 (59.1) |
| Epileptogenic zone localization, n (%) | *Temporal Plus* | 21 (70.0) | 16 (81.8) |
|  | *Prefrontal Plus* | 3 (10.0) | 1 (4.5) |
|  | *Posterior* | 3 (10.0) | 2 (9.1) |
|  | *Insulo-opercular* | 2 (6.7) | 1 (4.5) |
|  | *Centra-premotor* | 1 (3.3) | 0 (0.0) |
| MRI-visible lesion, n (%) | *3T* | 8 (26.7) | 6 (27.3) |
|  | *7T* | 8 (26.7) | 6 (27.3) |
| SEEG performed, n (%) | - | 22 (73.3) | 22 (100) |
| Surgery performed, n (%) | - | 14 (46.7) | 11 (50.0) |
| Histological diagnosis, n (%) | *No lesion* | 1 (3.3) | 1 (4.5) |
|  | *FCD type 1* | 3 (10.0) | 3 (13.6) |
|  | *FCD type 2* | 3 (10.0) | 1 (4.5) |
|  | *FCD type 3* | 1 (3.3) | 1 (4.5) |
|  | *Mild gliosis* | 2 (6.7) | 2 (9.1) |
|  | *Ganglioglioma* | 1 (3.3) | 1 (4.5) |
|  | *Heterotopia* | 1 (3.3) | 1 (4.5) |
| Surgical outcome, n (%) | *Engel I* | 8 (26.7) | 6 (27.3) |
|  | *Engel II* | 5 (16.7) | 4 (18.2) |
|  | *Engel III* | 1 (3.3) | 1 (4.5) |
|  | *Engel IV* | 0 (0) | 0 (0) |

### Thalamic sodium homeostasis and functional integrity

Representative voxel-wise reconstructions of thalamic TSC and *f_Na_* are shown in Figs. 1A and B. Compared with controls, patients exhibited higher whole-thalamus TSC (*F*_1,48_ = 9.141, *p* = 0.004) and *f_Na_* (*F*_1,48_ = 8.164, *p* = 0.006) z-scores. Whereas TSC elevations were distributed across medial, lateral and posterior thalamic segments, increased *f_Na_* was confined to the lateral thalamus (*t*_48_ = 3.585, *p* = 0.003). Neither metric differed between ipsilateral and contralateral sides.

Resting-state functional MRI revealed a different spatial pattern. Whereas the fractional amplitude of low-frequency BOLD fluctuations (fALFF) did not differ between groups (Fig. 2C), regional homogeneity (ReHo) of the BOLD signal was increased across the whole thalamus (*F*_1,48_ = 4.881, *p* = 0.032), with the strongest effects observed in the posterior segment (Fig. 2D). As for the sodium metrics, fALFF and ReHo did not differ between ipsilateral and contralateral thalami.

**Fig. 2.**
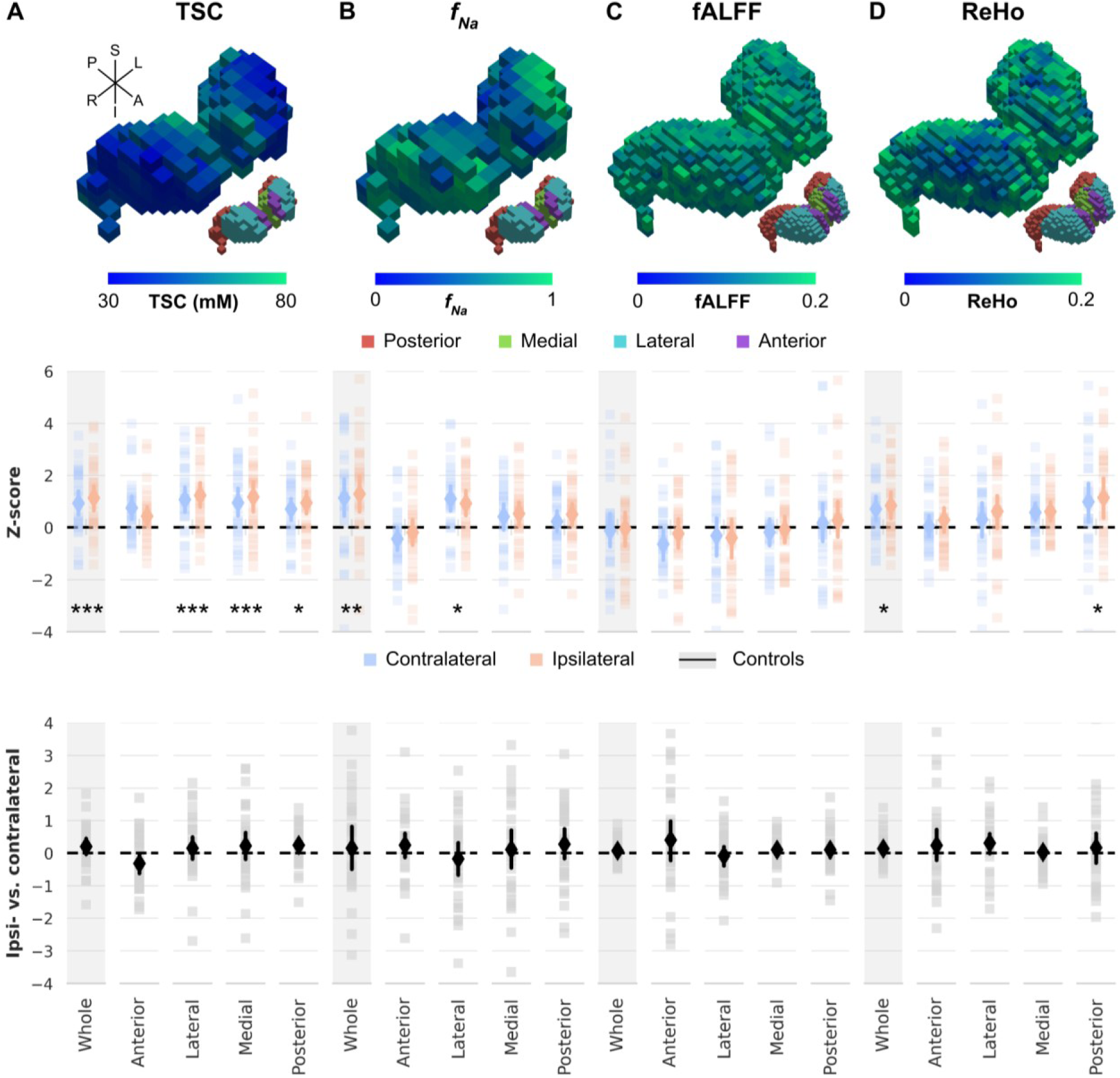
Thalamic sodium homeostasis and functional integrity in patients. (A) Example voxel-wise 3D reconstruction of thalamic total sodium concentration values (TSC, top), group-averaged whole thalamus and segment- and side-wise z-scores (middle), and ipsi-vs. contralateral differences. (B–D) Same format as (A), but for *f*_Na_, fractional amplitude of low-frequency fluctuations (fALFF), and regional homogeneity (ReHo), respectively. Solid lines indicate mean z-scores across patients and controls; shaded areas represent 95% confidence intervals. Statistical annotations \**p* < 0.05, \*\**p* < 0.01, \*\*\**p* < 0.005, and \*\*\*\**p* < 0.001 indicates group differences.

An overview of parameter-, segment-, and side-wise z-scores, as well as corresponding ipsi-vs. contralateral differences, with patients stratified by epilepsy type (i.e., TLE vs. NTLE) can be found in Supplementary Figs. 2 and 3.

### Multiparametric assessment

The observed alterations in sodium homeostasis and intrinsic function were next considered alongside previously reported structural metrics (volume and T_1_) to derive a multivariate thalamic MRI profile of focal epilepsy^13^. An overview of subject-wise z-scores across all parameters, segments, and sides is displayed in Supplementary Fig. 4.

#### Group differentiation

Mean-centered partial least squared (PLS) analyses identified a single latent variable (LV) differentiating patients from controls (*p_perm_* = 0.0002). Feature contributions, expressed by their bootstrapped ratio (BSR), varied across MRI parameters (*F*_5,38_ = 19.585, *p* < 0.0001) and thalamic segments (*F*_5,38_ = 7.175, *p* = 0.0006) with volumetric measures contributing most strongly, followed by TSC (all *p* < 0.05), whereas anterior thalamic features contributed least (Fig. 3A). To determine whether this thalamic MRI profile generalized to individual patients, features exceeding the significance threshold (|BSR| > 2.58; n = 14; Supplementary Table 1) were used to train multiple classifiers (Fig. 3B). Comparing receiver operator characteristic (ROC) curves, this feature set yielded significantly higher area under the curve (AUC) values across logistic regression (LR: 0.81 [CI: 0.68-0.95]) and support vector machine (SVM: 0.90 [CI: 0.81-0.99]) models compared with models trained on an equal number of low-contributing features (DeLong test, *p* < 0.0001 for both). Although classification performance was also higher for the Random Forest model, this difference did not reach statistical significance. Together, these findings indicate that multivariate PLS-derived feature ranking robustly identifies MRI markers with meaningful individual-level discriminative power across multiple classifier architectures.

**Fig. 3.**
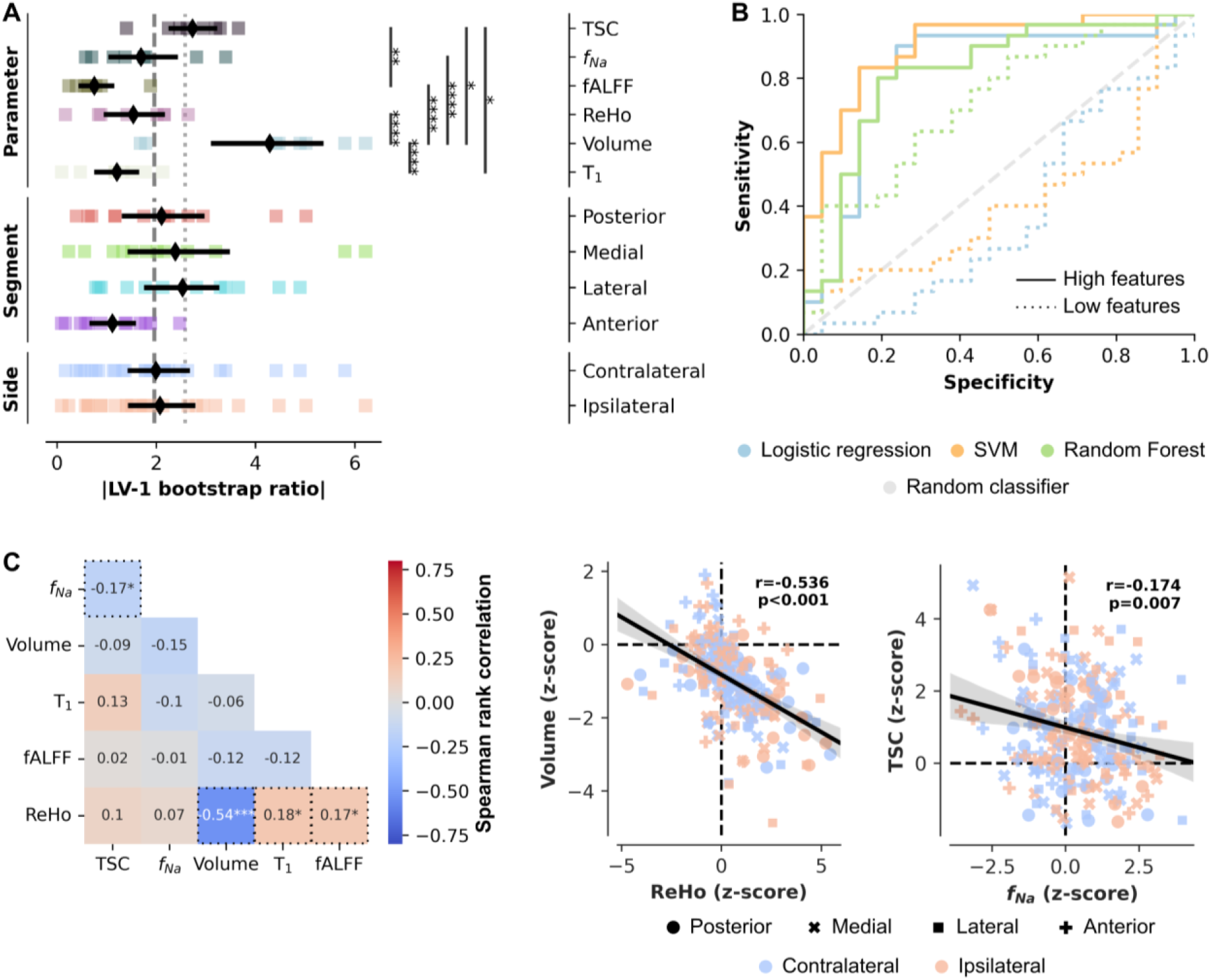
Integration of MRI features. (A) Strip plot showing bootstrapped ratios (BSRs) for individual MRI features contributing to the significant latent variable distinguishing patients from controls, as well as their averages (±95% CI) after grouping by parameter, segment, and side. (B) Receiver operating characteristic plot for the classification models using the high-contributing (BSR ≥ 2.58, solid lines) and low-contributing (dotted lines) MRI features. (C) Heatmap of Spearman rank correlation coefficients among MRI features (collapsed across all segments and sides) and two example scatterplots that show significant relationships. *p < .05, **p < .01, ***p < .005, ****p < .001.

#### Correlation and causality analysis

Having identified a multiparametric MRI profile of focal epilepsy, we next asked whether its constituent features represent independent abnormalities or coordinated manifestations of a common pathological process. Spearman rank correlations revealed several significant associations among MRI parameters (Fig. 3C). ReHo was negatively correlated with thalamic volume (*r* = -0.54, *p* < 0.0001) but positively correlated with T_1_ (*r* = 0.18, *p* = 0.026) and fALFF (*r* = 0.17, *p* = 0.026). Higher TSC values were inversely related to *f_Na_* (*r* = -0.17, *p* = 0.035), while thalamic atrophy was modestly associated with increases in *f_Na_* (*r* = -0.15, *p* = 0.052). Side-specific analyses (Supplementary Fig. 5) further showed greater ipsilateral-contralateral differences for correlations involving *f_Na_* in patients than in controls.

We then performed mediation analyses to examine whether the observed associations followed a hierarchical organization. Three significant indirect (bidirectional) effects were identified: ReHo statistically mediated the association between thalamic volume and T_1_ (Supplementary Fig. 6A), and the association between ReHo and volume was further linked to both *f_Na_* (Supplementary Fig. 6B) and TSC (Supplementary Fig. 6C). Together, these mediation pathways are consistent with an organization in which alterations in tissue structure and functional integrity might be coupled to changes in sodium homeostasis.

#### Electrophysiological and clinical correlation

The translational relevance of the MRI profile was subsequently evaluated by relating it to direct electrophysiological measurements obtained with SEEG. Twenty-two patients had SEEG electrodes implanted within at least part of the (predominantly ipsilateral) thalamus. A probabilistic coverage map illustrates the spatial distribution of thalamic bipolar contacts across patients (Fig. 4A). Electrode sampling was strongly concentrated within the posterior thalamus, where contact density was markedly higher than in the remainder of the thalamus (9.34 ± 6.52% vs. 0.46 ± 1.37%, p < 0.0001). Consequently, SEEG-derived measures were evaluated both across the entire ipsilateral thalamus and within the ipsilateral posterior segment.

**Fig. 4.**
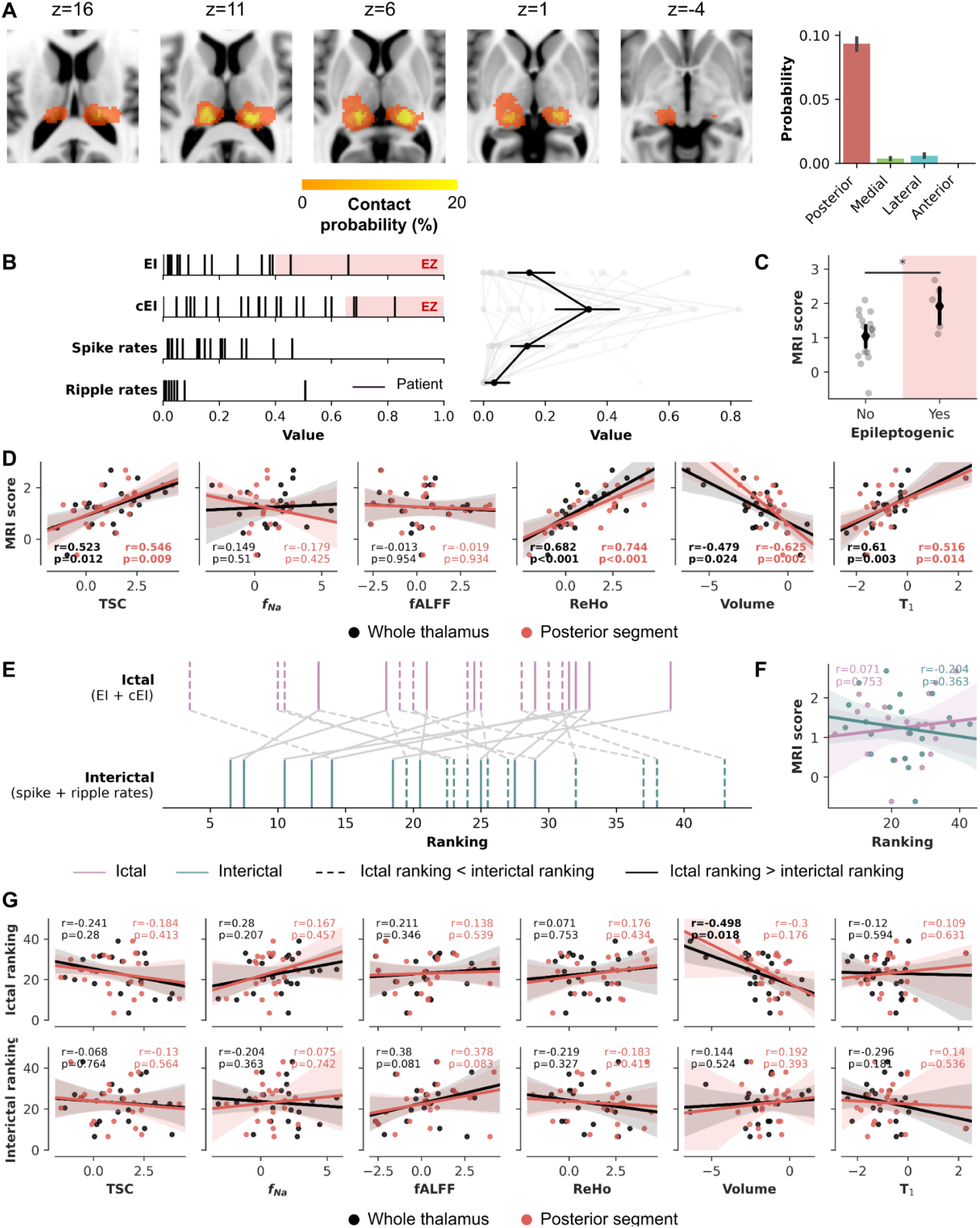
SEEG-derived electrophysiological correlates of thalamic MRI abnormalities. (A) Probabilistic contact map showing spatial distribution of SEEG bipolar electrode coverage within the thalamus, and the average sampling densities within segments. (B) Distribution of ictal (EI, cEI) and interictal (spike, ripple rates) indices across patients (black vertical lines) with sampled thalamic contacts. (C) Comparison of MRI scores between patients with and without epileptogenic thalami. (D) Correlation plots linking thalamic MRI parameters (whole thalamus and posterior segment) with MRI scores. (E) Patient-specific ictal and interictal ranking. (F) Correlation plots linking ictal (purple) and interictal (green) ranking and MRI scores. (G) Correlation plots linking thalamic MRI parameters (whole thalamus and posterior segment) with electrophysiological rankings (ictal and interictal).

#### Epileptogenicity indices

To quantify thalamic involvement in epileptic activity, we derived ictal and interictal electrophysiological markers from SEEG recordings using the EI and cEI, and spike and ripple rates. Five patients exhibited clear ictal thalamic recruitment based on either elevated normalized EI (≥0.4) and/or cEI values (≥0.65), while interictal markers such as normalized spike and ripple rates remained below 0.5 in the majority (Fig. 4B). These observations enabled evaluation of whether the MRI profile identified electrophysiologically active thalami. Patients with SEEG-defined epileptogenic thalami exhibited higher MRI (profile) scores, computed as a weighted combination of the highest-contributing ipsilateral MRI features (|BSR| > 2.58; Supplementary Table 1) than the remaining SEEG cohort (Mann-Whitney U = 16.0, *p* = 0.039, δ = -0.624; Fig. 4C). This difference disappeared when the profile was replaced by an equally sized set of low-contributing MRI features (U = 54.0, *p* = 0.401, δ = 0.271), indicating that the association appears specific to the disease-relevant multivariate signature. Higher MRI scores were strongly associated with higher TSC, increased ReHo, reduced volume and longer T_1_, and thus primarily captured pathological changes (Fig. 4D, all *|r|* > 0.500, *p* < 0.05).

Although threshold-based classification of EI and cEI is clinically useful for identifying epileptogenic thalami, it reduces thalamic involvement to a binary outcome. We therefore derived composite ictal (EI and cEI) and interictal (spike and ripple rate) rankings to provide continuous measures of thalamic electrophysiological involvement (Fig. 4E). This approach captures gradations in ictal recruitment while enabling assessment of whether interictal activity provides complementary information beyond that captured by clinically defined epileptogenicity. Despite distinguishing patients with and without epileptogenic thalami, the multivariate MRI (profile) score was not significantly associated with either the ictal or interictal ranking (Fig. 4F), suggesting that it primarily reflects the presence rather than the degree of thalamic epileptogenicity. Among the individual MRI features, however, higher ictal rankings were associated with greater thalamic atrophy (*r* = -0.498, *p* = 0.018), consistent with stronger structural compromise in patients exhibiting greater ictal recruitment (Fig. 4G). This relationship remained directionally consistent when restricted to the posterior thalamus, although effect sizes were attenuated, and was partially accounted for by the extent of the epileptogenic network (partial *r* = -0.281). Interictal rankings and the relative dominance of ictal versus interictal activity showed no systematic associations with MRI or electrophysiological measures (Supplementary Fig. 7).

#### Epileptogenic network characteristics

The final set of analyses examined how thalamic imaging and electrophysiological measures relate to variability in clinical phenotype across patients (Fig. 5). Ictal thalamic involvement was positively associated with the extent of the epileptogenic network, such that patients with higher ictal rankings exhibited a greater number of epileptogenic regions (*r* = 0.428, *p* = 0.047; Fig. 5A). In contrast, interictal thalamic activity was not significantly associated with network extent or other clinical measures.

**Fig. 5.**
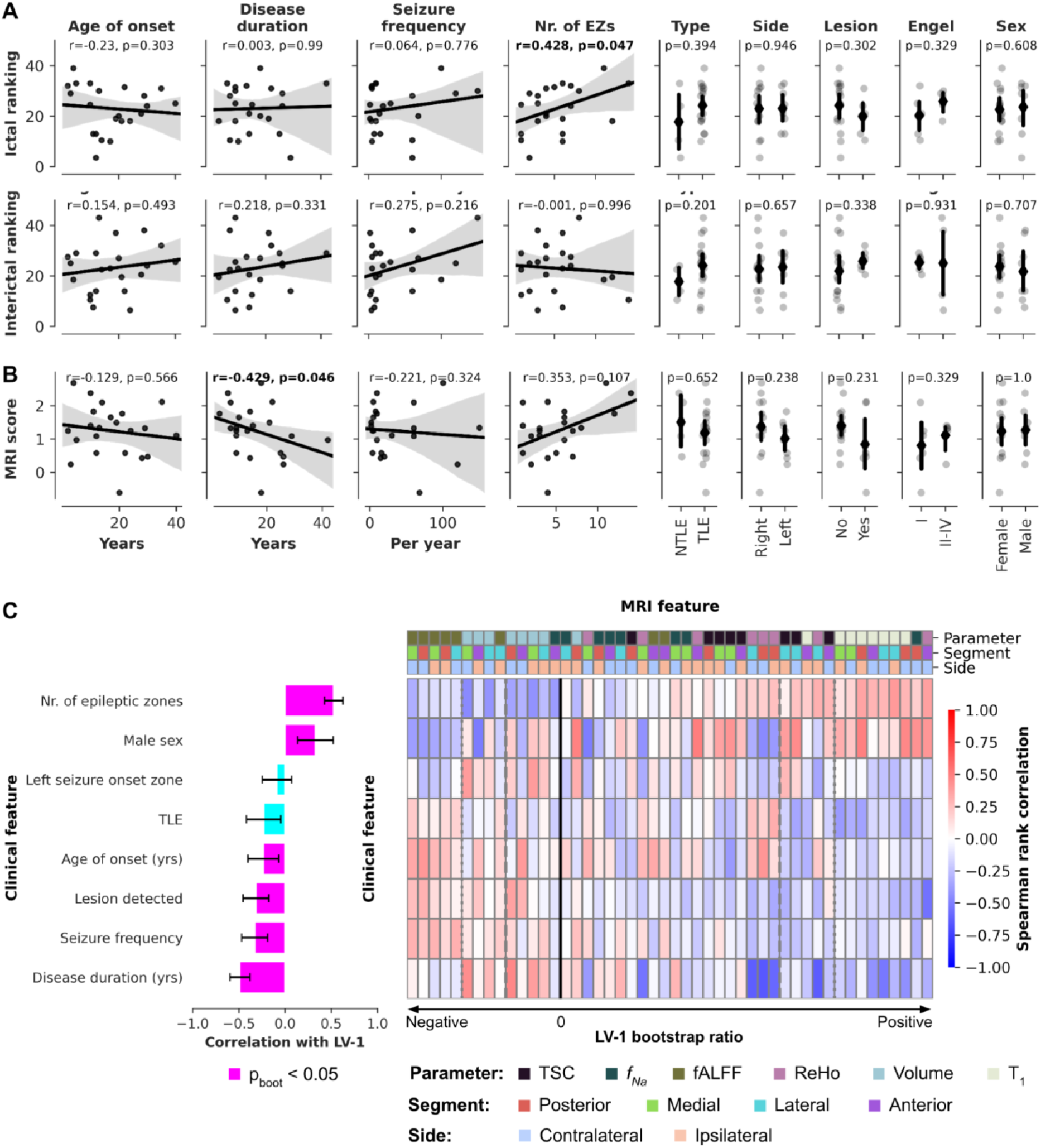
Clinical relevance of thalamic electrophysiological and MRI alterations. Scatterplots showing associations between clinical features and (A) ictal and interictal SEEG rankings, and (B) MRI score. (C) Behavioral partial least squares (PLS) analysis: dominant latent variable (LV-1) linking clinical features to MRI metrics. The bar plot represents the correlations between each clinical feature and the latent variable whereas the heatmap shows the correlation between clinical and MRI features. MRI and clinical features are ranked by bootstrapped ratios. Color coding corresponds to MRI parameter, segment, and laterality (as in Fig. 3).

We next asked whether the multivariate MRI profile derived from the patient-control analysis similarly captures clinical heterogeneity (Fig. 5B). Overall, subject-specific MRI (profile) scores showed only limited associations with clinical characteristics. The only significant relationship was observed for disease duration, with stronger expression of the MRI profile in patients with shorter disease duration (*r* = 0.420, *p* = 0.046), whereas the association with epileptogenic network extent did not reach significance (*r* = -0.353, *p* = 0.10). Together, these findings suggest that the patient-control profile primarily captures a common disease-related imaging phenotype rather than the clinical heterogeneity observed across patients.

Because the mean-centered PLS was designed to maximize patient-control separation rather than explain inter-individual clinical variability, we applied behavioral PLS to identify MRI features specifically associated with differences in clinical phenotype. Behavioral PLS identified two significant latent variables (Supplementary Fig. 6). The dominant LV (42.8% explained variance, *p_perm_* = 0.001) revealed two opposing MRI patterns associated with distinct aspects of the clinical phenotype. Volume and fALFF decrease with increasing numbers of epileptogenic regions while increasing with longer disease duration. In contrast, T_1_ and ReHo exhibited the opposite pattern, increasing with network extent but decreasing with disease duration (Fig. 5C). The second LV (Supplementary Fig. 6B) captured a complementary demographic dimension, with ReHo increasing with later disease onset and decreasing in males, whereas thalamic volume showed the opposite associations.

## Discussion

By combining multiparametric MRI with intracranial electrophysiology, we identified a profile of thalamic pathology in patients with DRFE. Rather than describing isolated structural, functional or ionic abnormalities, this framework captures complementary dimensions of thalamic pathology and relates them to electrophysiological involvement and clinical network organization. Together, our findings support the view that thalamic pathology reflects coordinated alterations across tissue structure, sodium homeostasis and intrinsic function, consistent with the thalamus acting as a pathological node within epileptogenic networks.

### Characterization of sodium homeostasis, structural, and functional abnormalities

Across patients, we observed a robust and global increase in thalamic TSC. Although atrophy-related spilling effects could theoretically elevate sodium measurements, the accompanying reductions in T_1_ and our gray matter density-based partial volume correction argue against this being the primary explanation. This widespread elevation is thus most consistent with diffuse impairment in ionic homeostasis and may reflect a combination of altered Na⁺/K⁺-ATPase function^58^, changes in intracellular-extracellular sodium gradients^20^, and/or glial responses to persistent network disruption^59^. By contrast, *f_Na_* increases were more spatially circumscribed and most pronounced in lateral thalamic regions. This pattern parallels prior cortical findings in DRFE patients, in which TSC elevations are diffuse but *f_Na_* abnormalities occur in sites characterized by hyperexcitability^23,60^. These dissociable sodium signatures may index distinct underlying processes: global metabolic stress versus localized disruptions in neuronal electrophysiology.

Resting-state fMRI revealed complementary alterations in intrinsic thalamic function. ReHo and fALFF capture distinct aspects of BOLD signal organization: ReHo quantifies the temporal synchrony of BOLD fluctuations across neighboring voxels and is commonly interpreted as an index of local functional coordination, whereas fALFF reflects the relative contribution of low-frequency oscillations and is thought to index the magnitude of intrinsic functional activity. fALFF did not differ systematically between patients and controls. In contrast, we observed increased ReHo, most prominently in posterior thalamic nuclei, suggesting abnormal local synchrony within thalamocortical circuits, potentially reflecting enhanced or maladaptive coordination^61,62^. Given the thalamus’ role as a highly connected hub capable of amplifying and distributing brain activity^63^, these functional changes may facilitate the propagation of epileptic seizures across distributed networks^64^.

A key finding of this study is that complementary MRI measures jointly define a coherent profile of thalamic pathology. Whereas previous work has primarily interpreted structural, functional and sodium abnormalities independently, our multivariate analyses demonstrate that these measures provide complementary rather than redundant information. The resulting profile robustly distinguished patients from controls, generalized across multiple machine-learning classifiers, and was further linked to electrophysiological measures of thalamic involvement. When assessed jointly, volume and TSC emerged as the dominant contributors to a latent pattern, which not only highlights structural degeneration and impaired sodium regulation as key thalamic markers of DRFE-related pathology, but also allowed accurate automatic classification of patients and controls, with performance comparable to previous reports^65^. In line with our previous structural 7T MRI study, the strongest changes in sodium and functional MRI features tend to predominate in lateral, medial and posterior nuclei rather than anterior nuclei^13^. While this reduced anterior contribution appears reproducible, it should be interpreted with caution: the lower spatial resolution of ^23^Na MRI may limit sensitivity to small nuclei and could therefore partially influence the relative weighting of features in the PLS analysis. Nonetheless, the overall pattern reinforces the importance of a nucleus-aware, multiparametric characterization of thalamic pathology, rather than treating the thalamus as a homogeneous structure^3,66^.

Importantly, the identified profile was not dominated by a single imaging modality but emerged from the complementary contribution of structural, functional and sodium-sensitive MRI measures. This underscores that thalamic pathology is inherently multidimensional, with different MRI contrasts capturing distinct but interrelated aspects of tissue integrity. Indeed, the observed relationships between MRI measures suggest that these abnormalities are not independent but instead reflect coordinated pathological processes. In particular, thalamic atrophy was strongly associated with increased ReHo, suggesting that structural degeneration may either promote or arise from heightened local functional synchrony. This structural-functional link is not surprising as alterations in resting-state fMRI metrics, including ReHo, have been widely documented in clinical populations with structural degeneration^67^. The positive association between ReHo and fALFF further points to intrinsic reorganization of thalamic dynamics: patients with greater local synchrony also exhibited stronger low-frequency BOLD fluctuations, indicating that alterations in the coordination of neural activity may be accompanied by increased intrinsic oscillatory amplitude within compromised tissue. This relationship emerged despite the absence of group-level fALFF differences, suggesting that intrinsic functional reorganization is heterogeneous across patients and may not be captured by univariate group comparisons alone.

The sodium-sensitive MRI measures further reinforced the multidimensional organization. The inverse association between TSC and *f_N_*_a,_ together with their distinct spatial distributions, supports the hypothesis that these metrics characterize different aspects of sodium physiology^28^. TSC may reflect diffuse ionic imbalance or glial dysfunction, whereas *f_Na_* may be more sensitive to localized sodium alterations linked to neuronal hyperexcitability^23,60^. Notably, correlations involving *f_Na_* differed more strongly between ipsilateral and contralateral thalami in patients than in controls, suggesting lateralized sodium dysregulation related to the EZN. This hemispheric asymmetry reinforces the potential relevance of *f_Na_* as a marker of focal thalamic involvement within epileptogenic networks.

Together, the mediation analyses suggest that these structural, functional, and ionic abnormalities should not be interpreted as isolated imaging findings but rather as interconnected components of a common thalamic pathological phenotype. Although causal inference is beyond the scope of this cross-sectional study, the identified indirect associations provide a first framework for understanding how complementary MRI markers may interact to characterize pathological network nodes. Rather than replacing individual MRI measures, this integrative perspective highlights the value of combining complementary contrasts to capture different facets of thalamic pathology.

### Bringing together MRI, SEEG and clinical features

By bridging imaging and intracranial electrophysiology, we found that the multiparametric MRI profile identified patients with electrophysiologically defined epileptogenic thalami. Although the profile was derived solely from patient-control differences, it generalized to a clinically distinct comparison, with patients showing ictal thalamic involvement exhibiting significantly stronger expression of the disease profile than those without thalamic epileptogenicity. Importantly, this association was not observed for a control score derived from low-contributing MRI features, supporting the biological specificity of the identified profile rather than an arbitrary combination of imaging abnormalities.

Examining the individual imaging components underlying this profile further showed that reduced thalamic volume was consistently associated with greater ictal epileptogenicity. Patients with stronger thalamic recruitment during seizures exhibited more pronounced atrophy, consistent with growing evidence that the thalamus acts as an active node in seizure propagation rather than a passive relay^2,9,10^. This association may reflect cumulative structural effects of repeated ictal recruitment or, conversely, increased vulnerability of structurally compromised thalamic nuclei to seizure generation. The consistency of this relationship across whole-thalamus and posterior-segment analyses further supports its robustness.

Beyond thalamic epileptogenicity itself, greater ictal thalamic recruitment was associated with a more extensive epileptogenic network, supporting previous observations that thalamic involvement scales with network complexity and spatial seizure propagation^2^. Interestingly, however, the patient-control MRI score showed only limited relationships with broader clinical heterogeneity, suggesting that it primarily captures disease-related abnormalities shared across patients. Behavioral PLS addressed this complementary question by identifying MRI features associated with inter-individual clinical variability. Whereas structural and functional MRI measures were differentially associated with epileptogenic network extent, disease chronicity, and demographic characteristics, the multivariate profile remained the strongest discriminator of electrophysiologically defined thalamic involvement. Together, these findings indicate that complementary multivariate models capture different aspects of disease: one describing the common pathological signature of focal epilepsy, the other explaining how this pathology varies across individuals.

### Toward precision imaging of thalamic pathology

A central implication of this study is that thalamic pathology is best understood as a multidimensional phenomenon rather than a collection of isolated structural, functional, or electrophysiological abnormalities. By integrating complementary MRI contrasts with intracranial electrophysiology, our framework links tissue-level alterations to network-level dysfunction, providing a more comprehensive characterization of the thalamus as a pathological network node in focal epilepsy. Within this framework, the thalamus appears particularly vulnerable to repeated recruitment during seizures. Its extensive structural and functional connectivity may facilitate seizure propagation while simultaneously predisposing it to cumulative structural remodeling, sodium dysregulation, and intrinsic functional reorganization. This interpretation is consistent with both human and experimental studies demonstrating secondary thalamic injury and altered thalamocortical circuitry in acquired epilepsies^9,13,16^.

More broadly, these findings illustrate how multiparametric imaging could support precision medicine in focal epilepsy. Current therapeutic decision-making, including epilepsy surgery and DBS, relies largely on anatomical landmarks and electrophysiological recordings despite substantial heterogeneity in thalamic pathology across patients. The multimodal framework presented here offers an alternative strategy by integrating complementary structural, functional, ionic homeostatic and electrophysiological information to characterize individual thalamic pathology. Such imaging profiles could ultimately support patient stratification, target selection and longitudinal monitoring of therapeutic response, complementing existing electrophysiological approaches^5,68,69^.

### Study limitations and future directions

Several limitations should be noted. First, despite segment-wise analysis, the spatial resolution of ²³Na MRI may not resolve small thalamic nuclei with complete accuracy. Future studies further leveraging ultra-high-field (7T) MRI and state-of-the-art imaging technologies may improve spatial specificity of TSC and *f_Na_* maps. Second, causality between the different MRI and electrophysiological features cannot be definitively established in cross-sectional data. Longitudinal studies ideally before and after therapeutic interventions such as DBS will be needed to clarify temporal dynamics. Third, although our SEEG-based epileptogenicity indices provide valuable insight, they reflect a subset of thalamic nuclei accessible to electrode implantation and may not capture the full extent of thalamic involvement.

Beyond focal epilepsy, the present framework illustrates how complementary imaging contrasts and electrophysiological recordings can be integrated to characterize pathological network nodes. Because pathological network nodes are a common feature of many neurological disorders, similar multimodal approaches may provide a general framework for identifying biologically meaningful imaging profiles across diseases.

## Data Availability

The data are not publicly available due to sensitive information that could compromise the privacy of research participants. Nonetheless, anonymized data are available from the corresponding author on reasonable request.

## Data and code availability

Code to perform the analyses and visualize the results used in this work can be found online (https://github.com/royhaast/smk-epilepsy-7T-sodium). The data are not publicly available due to sensitive information that could compromise the privacy of research participants. Nonetheless, anonymized data are available from the corresponding author on reasonable request.

## Acknowledgments

We are grateful to the patients and control participants who agreed to take part in this study. In addition, we would like to thank Patrick Viout, Lauriane Pini, Claire Costes, and Veronique Gimenez for data acquisition and study logistics. We would also like to express our gratitude to Shailesh Appukuttan for advice on the classification analyses.

## Supplementary Material

**Supplementary Fig. 1.**
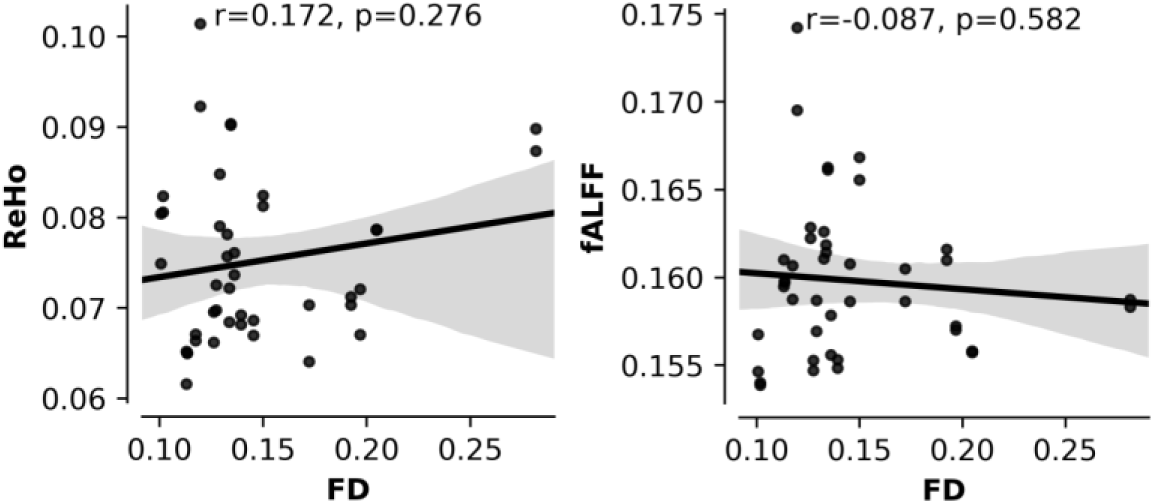
Assessment of head-motion effects on resting-state functional MRI measures. Scatterplots showing the relationship between framewise displacement (FD) and regional homogeneity (ReHo; left) and fractional amplitude of low-frequency fluctuations (fALFF; right) across participants. These analyses were performed to assess the sensitivity of the functional MRI metrics to head motion.

**Supplementary Fig. 2.**
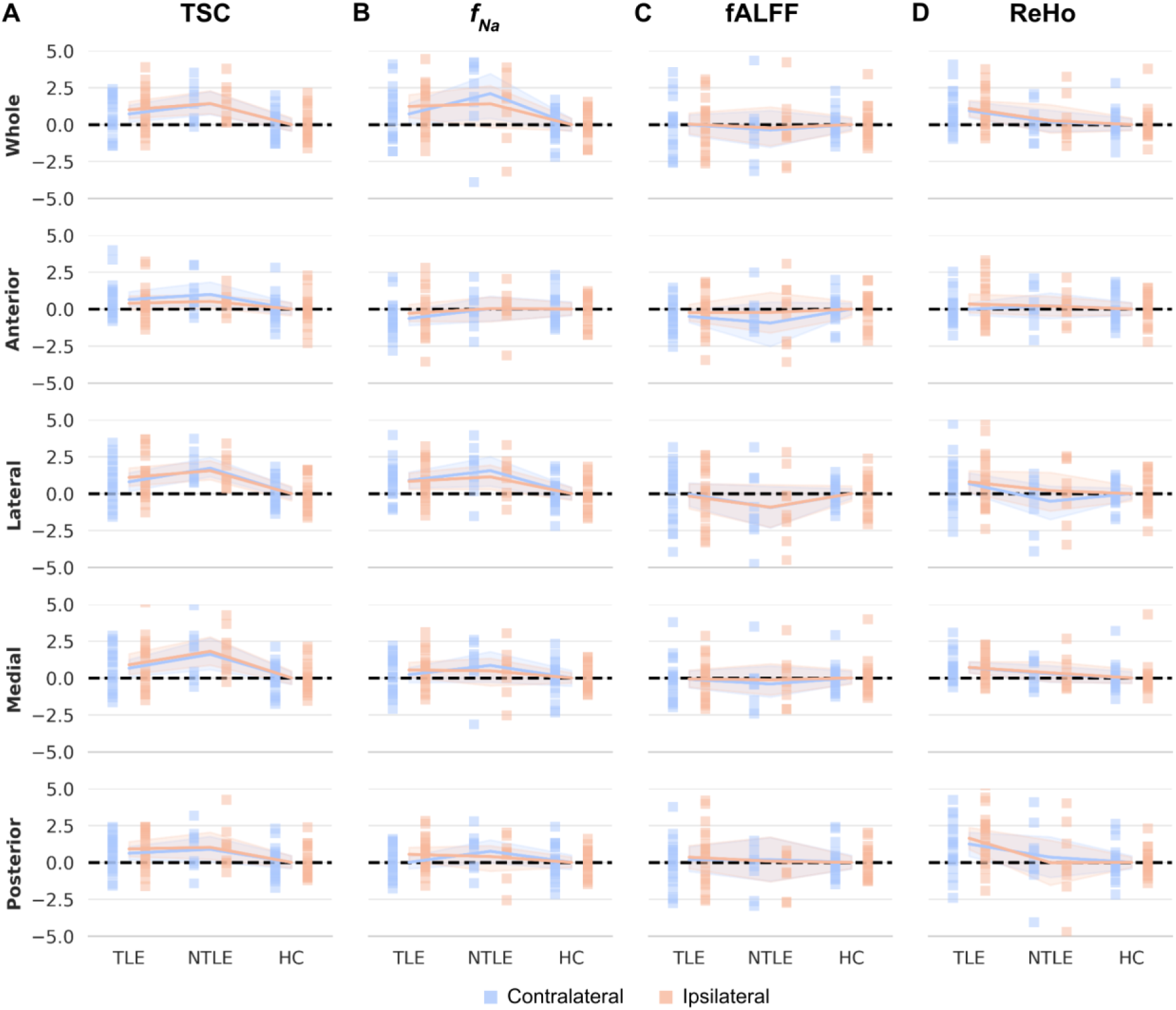
Group comparison of thalamic sodium homeostasis and intrinsic functional MRI measures. Whole-thalamus and segment-wise z-scores for (A) total sodium concentration (TSC), (B) short T_2_* sodium signal fraction (*f_Na_*), (C) fractional amplitude of low-frequency fluctuations (fALFF), and (D) regional homogeneity (ReHo) in patients with temporal lobe epilepsy (TLE), non-temporal lobe epilepsy (NTLE), and healthy controls (HC). Solid lines indicate group means; shaded areas represent 95% confidence intervals.

**Supplementary Fig. 3.**
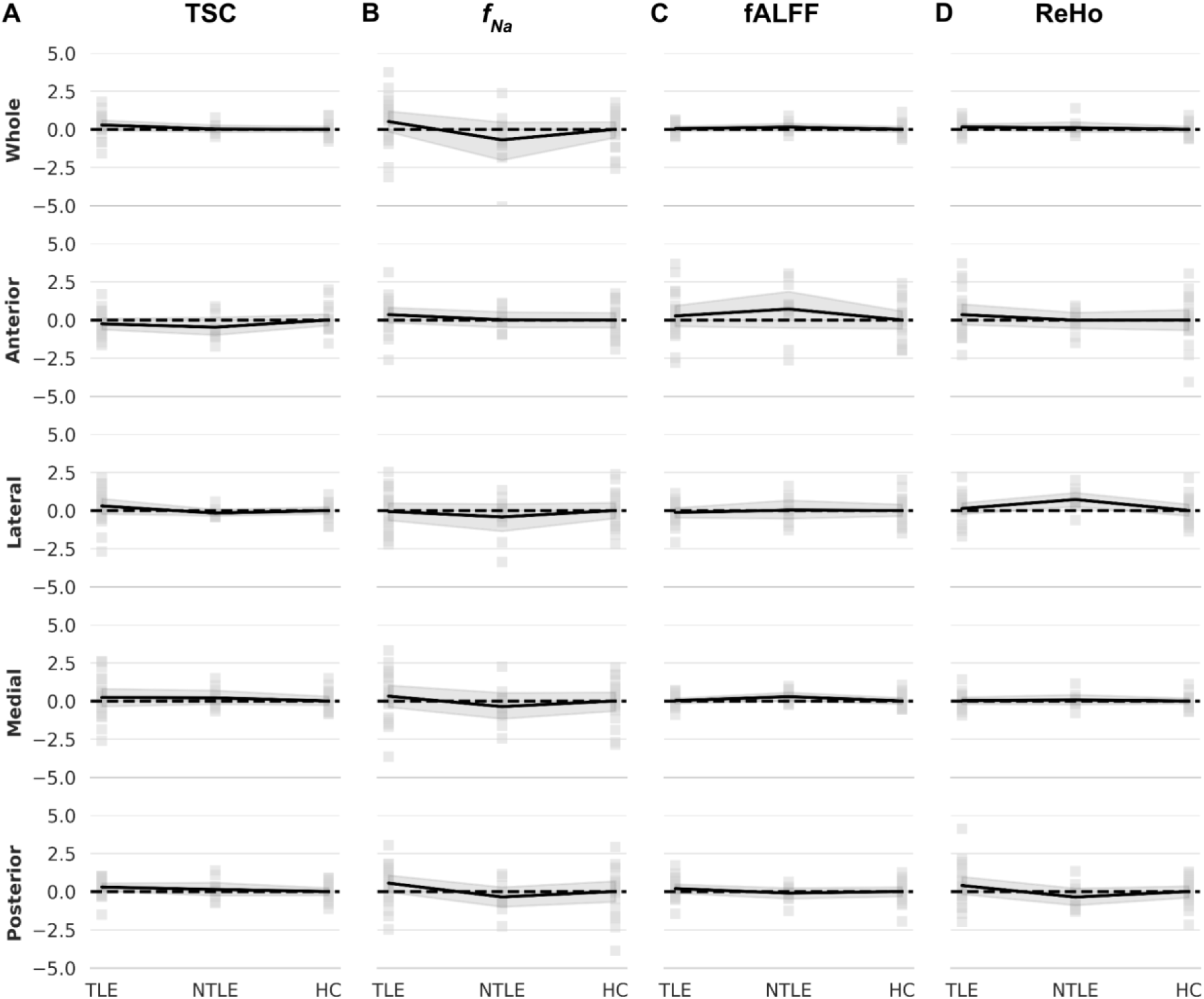
Ipsilateral versus contralateral comparison of thalamic sodium and functional MRI measures. Ipsilateral vs. contralateral z-scores for (A) total sodium concentration (TSC), (B) short T_2_* sodium signal fraction (*f_Na_*), (C) fractional amplitude of low-frequency fluctuations (fALFF), and (D) regional homogeneity (ReHo) across the whole thalamus and individual thalamic segments in patients with temporal lobe epilepsy (TLE), non-temporal lobe epilepsy (NTLE), and healthy controls (HC). For controls, hemispheres correspond to the left and right thalamus.

**Supplementary Fig. 4.**
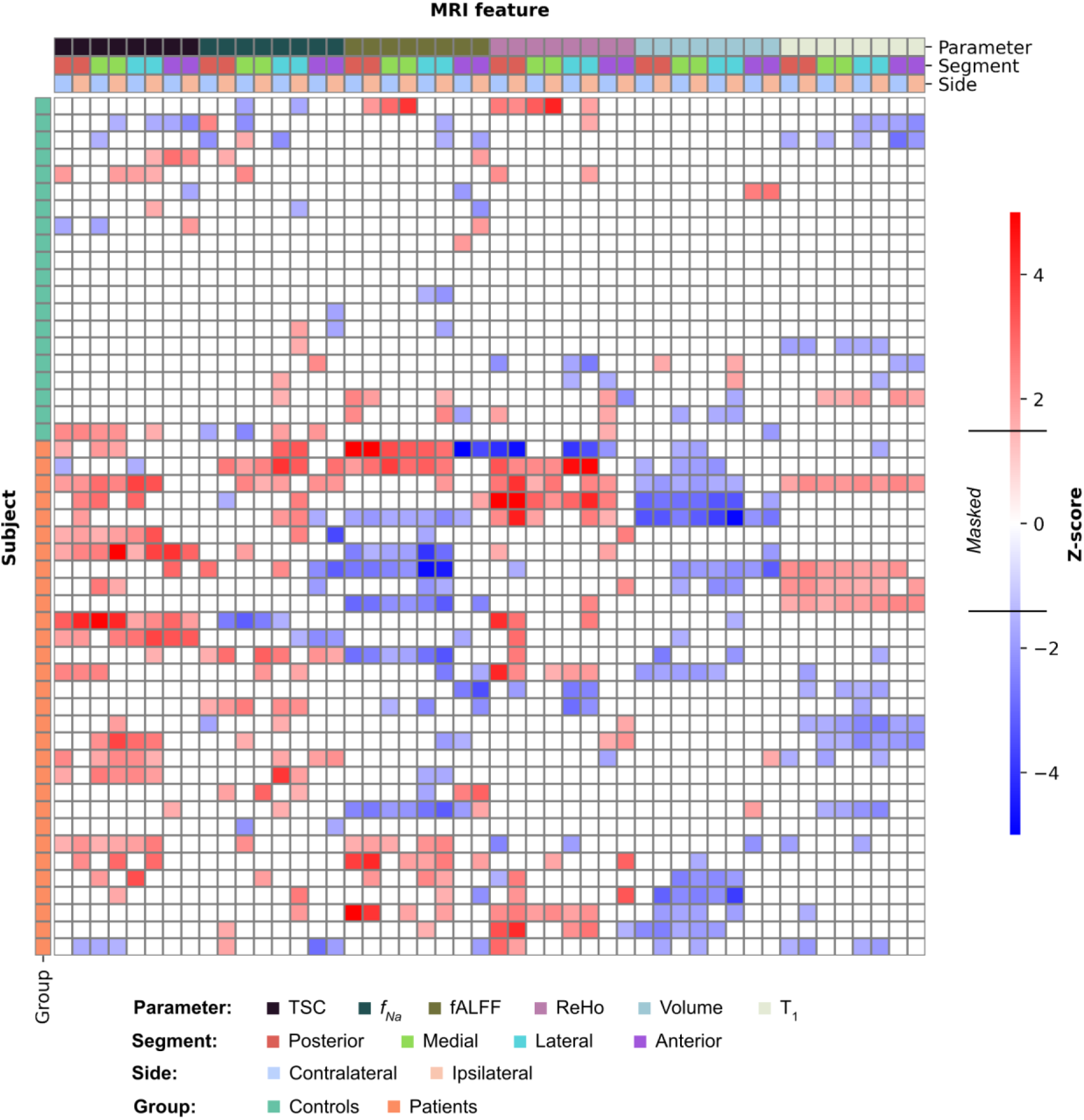
Z-score heatmap of individual thalamic MRI features. Color-coded matrix of MRI features across participants, segmented by metric (TSC, f_Na_, fALFF, ReHo, volume, T_1_), thalamic region (posterior, medial, lateral, anterior), and laterality (ipsilateral vs. contralateral to seizure onset in patients; left vs. right in controls). Values are z-scored and thresholded at ±1.5. Vertical bars on the left indicate group identity (controls vs. patients); horizontal bars above indicate feature type.

**Supplementary Fig. 5.**
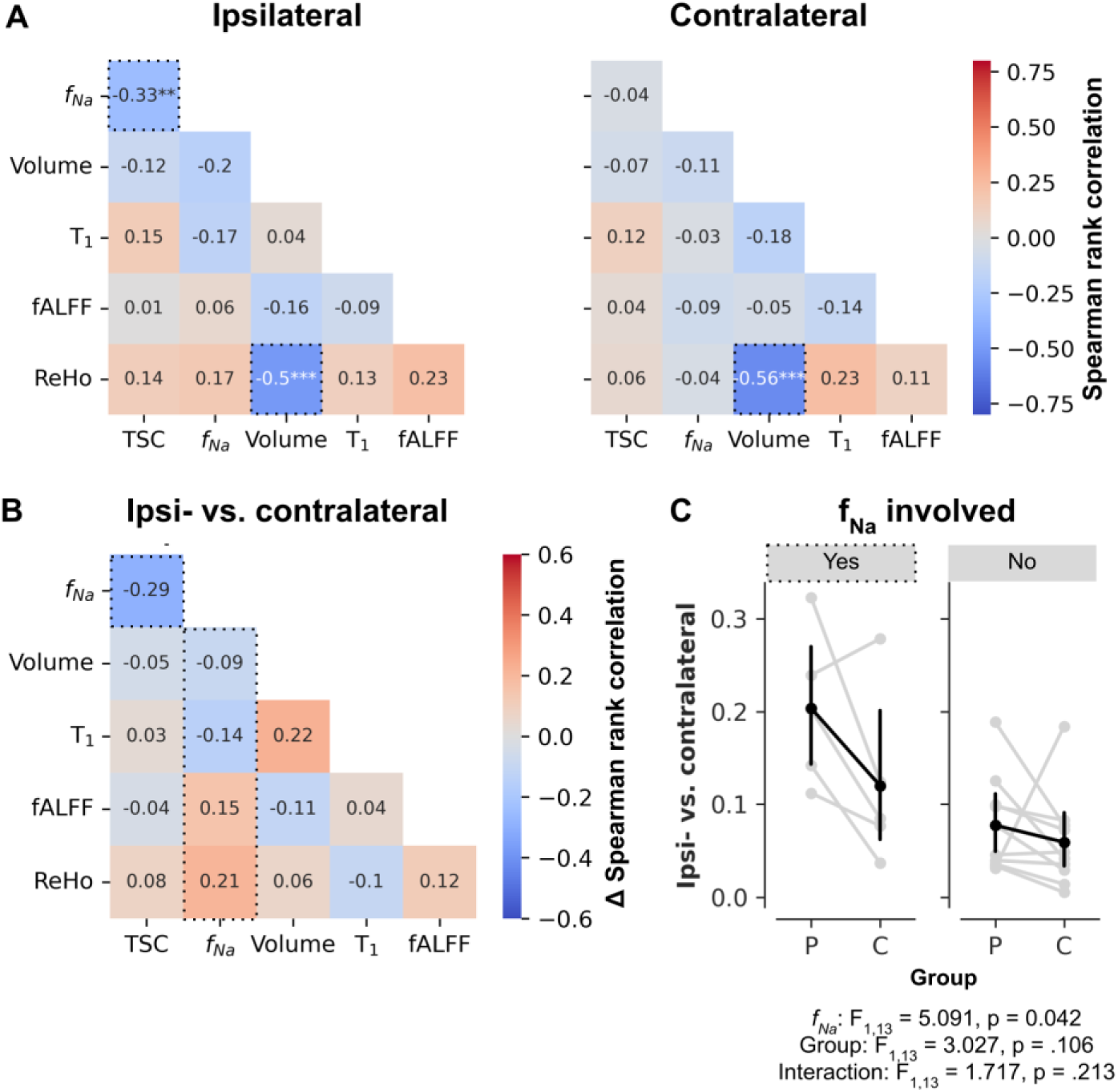
Side-specific correlations between thalamic MRI features. Spearman correlation matrices computed separately for (A) ipsilateral, contralateral, (B) and ipsi-vs. contralateral thalamic MRI features across patients. (C) Differences in correlation coefficients between comparisons involving *f_Na_* and those without *f_Na_*, illustrating the greater lateralization of *f_Na_*-related associations.

**Supplementary Fig. 6.**
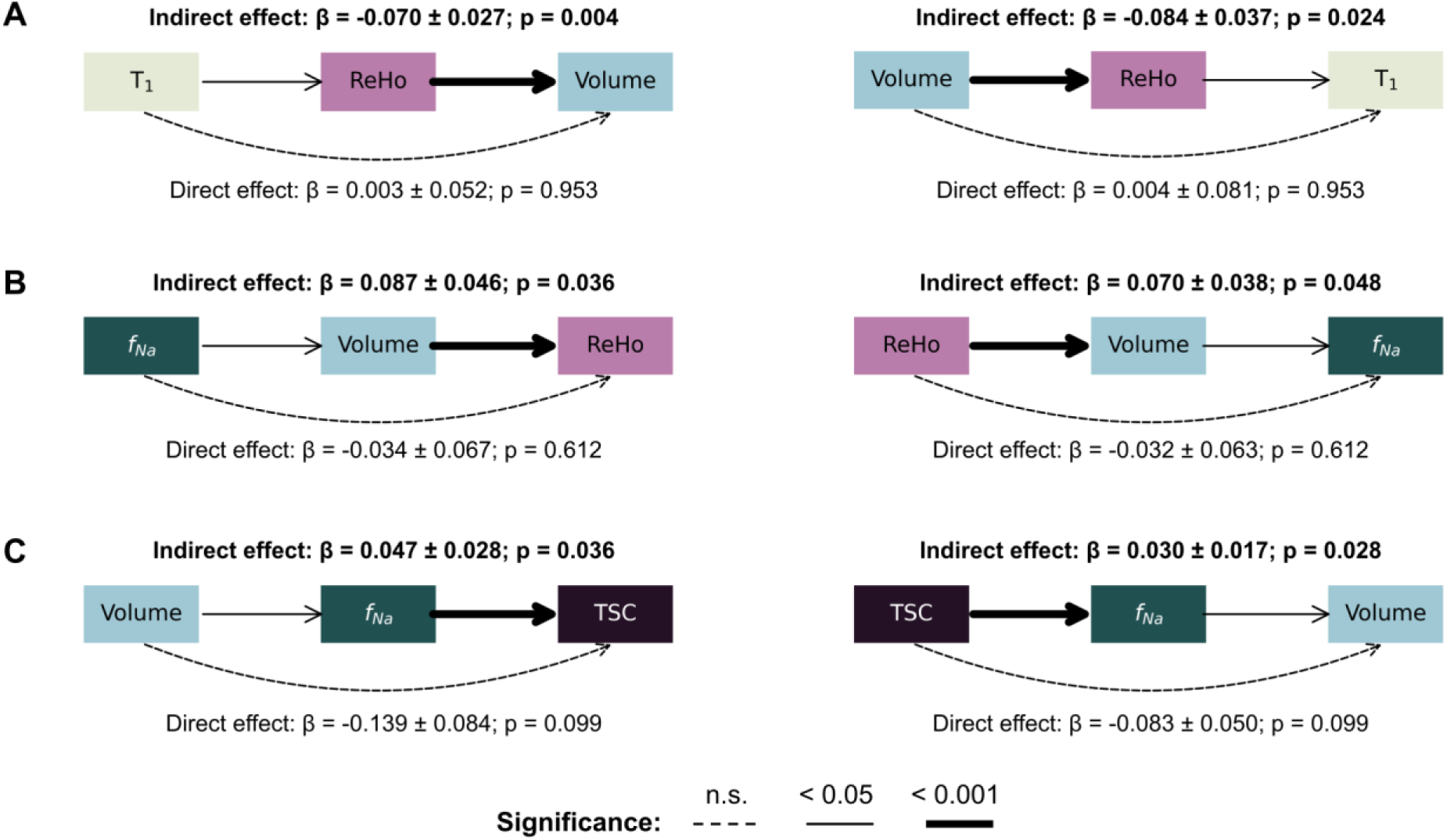
Mediation analyses linking structural, functional, and sodium-sensitive MRI measures. Significant mediation models identified from the pairwise MRI analyses. (A) Relationship between thalamic volume and T_1_ mediated by regional homogeneity (ReHo). (B) Relationship between the short T_2_* sodium signal fraction (*f_Na_*) and ReHo mediated by thalamic volume. (C) Relationship between thalamic volume and total sodium concentration (TSC) mediated by ReHo. Standardized regression coefficients are shown for each path; significant indirect effects were determined using bootstrap resampling.

**Supplementary Fig. 7.**
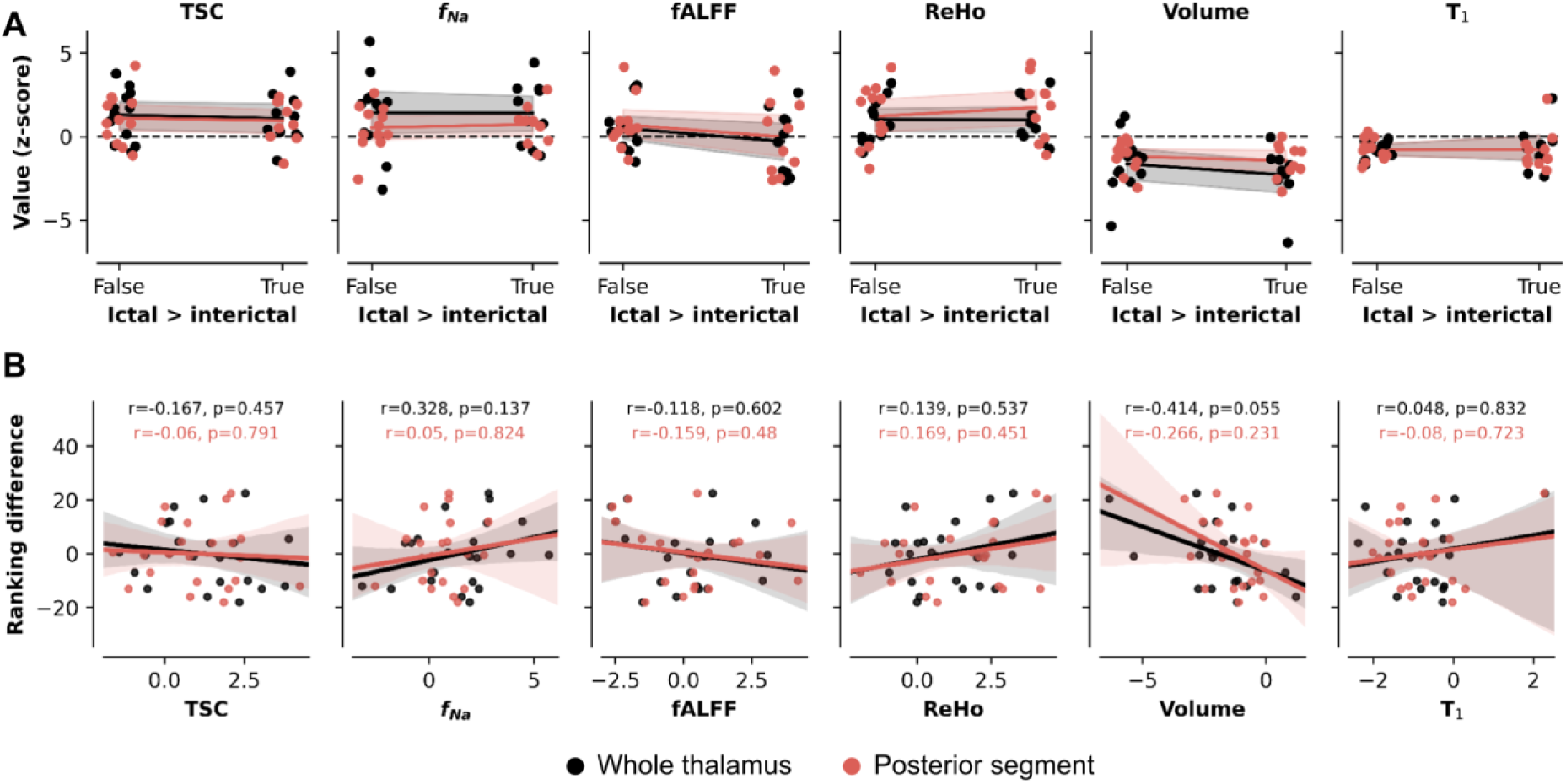
MRI features in relation to ictal and interictal thalamic involvement. (A) Comparison of MRI feature z-scores between patients with higher ictal than interictal electrophysiological rankings and those showing the opposite pattern. (B) Associations between MRI feature z-scores and the difference between ictal and interictal rankings across patients.

**Supplementary Fig. 8.**
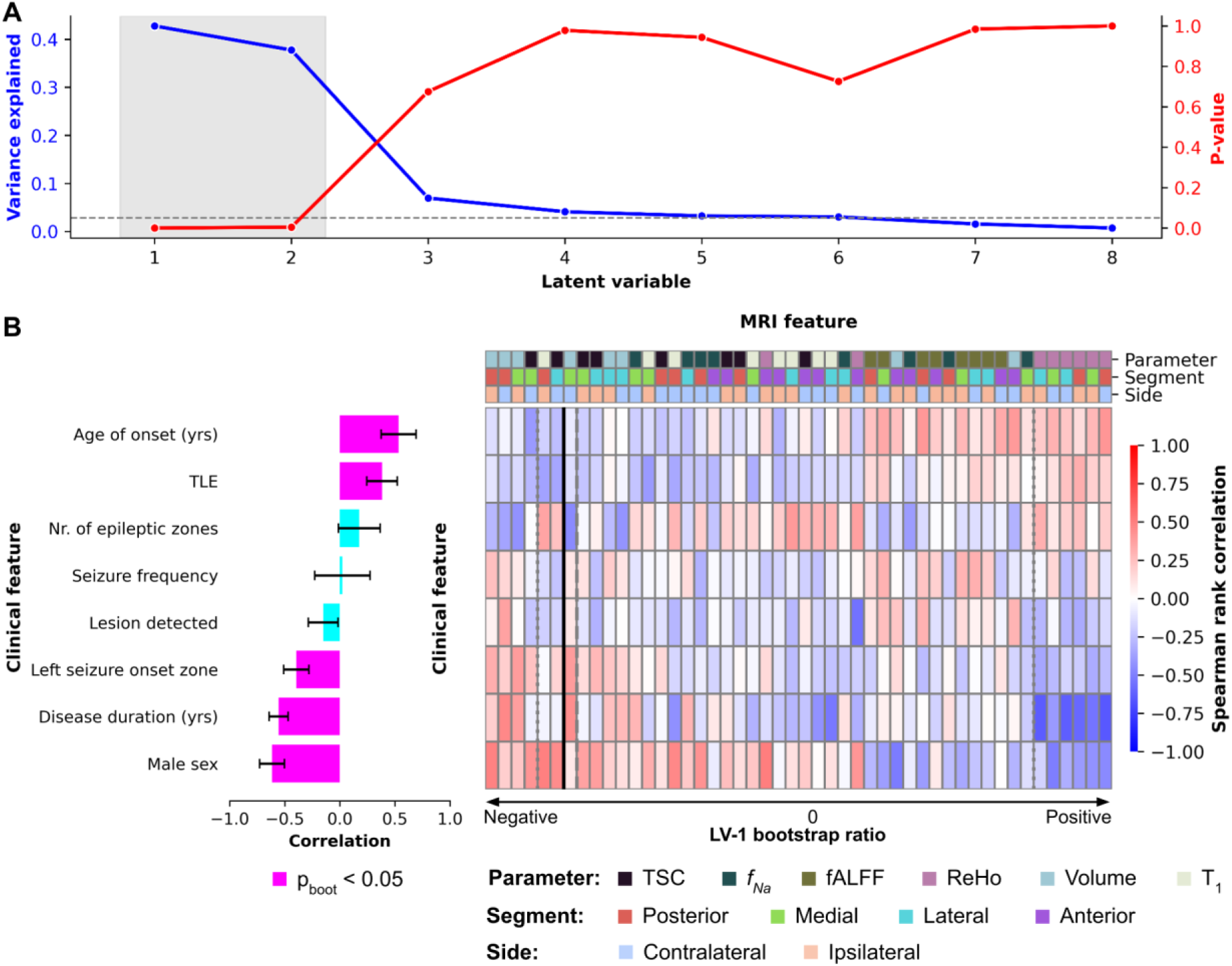
Behavioral partial least squares (PLS) analysis. (A) Percentage of explained covariance and corresponding permutation-derived p-values for each latent variable identified by behavioral PLS. (B) Feature loadings for the second significant latent variable (LV-2), showing the MRI and clinical variables contributing to this dimension of MRI-clinical covariation.

**Supplementary Table 1.** Overview of the MRI features with highest and lowest BSRs used as feature sets to the classifiers and composite MRI score.

| High-contributing features |  |  |  |
| --- | --- | --- | --- |
| *Parameter* | *Segment* | *Side* | *BSR* |
| Volume | Medial | Ipsilateral | 6.22 |
| Volume | Medial | Contralateral | 5.80 |
| Volume | Posterior | Ipsilateral | 5.02 |
| Volume | Lateral | Contralateral | 4.89 |
| Volume | Lateral | Ipsilateral | 4.47 |
| Volume | Posterior | Contralateral | 4.41 |
| TSC | Lateral | Ipsilateral | -3.65 |
| $f_{Na}$ | Lateral | Contralateral | -3.40 |
| TSC | Lateral | Contralateral | -3.30 |
| TSC | Medial | Ipsilateral | -3.20 |
| TSC | Posterior | Ipsilateral | -2.95 |
| $f_{Na}$ | Lateral | Ipsilateral | -2.81 |
| *ReHo* | Posterior | Ipsilateral | -2.64 |
| TSC | Medial | Contralateral | -2.63 |

| Low-contributing features |  |  |  |
| --- | --- | --- | --- |
| *Parameter* | *Segment* | *Side* | *BSR* |
| $T_1$ | Anterior | Ipsilateral | 0.10 |
| *ReHo* | Anterior | Contralateral | -0.18 |
| $fALFF$ | Medial | Ipsilateral | 0.24 |
| $fALFF$ | Posterior | Contralateral | -0.39 |
| $T_1$ | Anterior | Contralateral | -0.47 |
| $fALFF$ | Medial | Contralateral | 0.56 |
| $f_{Na}$ | Anterior | Ipsilateral | 0.58 |
| $fALFF$ | Anterior | Ipsilateral | 0.63 |
| $fALFF$ | Posterior | Ipsilateral | -0.63 |
| $f_{Na}$ | Posterior | Contralateral | -0.70 |
| $fALFF$ | Lateral | Contralateral | 0.78 |
| *ReHo* | Lateral | Contralateral | -0.84 |
| *ReHo* | Anterior | Ipsilateral | -0.89 |
| $fALFF$ | Lateral | Ipsilateral | 0.89 |

